# ‘*Certainly not in the heterosexual public eye*’: a qualitative study of the sociodemographic, behavioural and attitudinal drivers of syphilis among heterosexual-identifying people in England (the NEXUS study)

**DOI:** 10.1101/2025.05.06.25327060

**Authors:** Megan Walsh, David Reid, Josh Forde, Lynsey Emmett, Medhat Basta, David Phillips, Malini Raychaudhuri, Freddy Green, Will Nutland, Alison R Howarth, Mateo Prochazka, Gwenda Hughes, Kirsty Foster, Catherine H Mercer, Hamish Mohammed

## Abstract

**Background:** Recent increases in syphilis diagnoses among heterosexual individuals are a growing public health concern. We aimed to qualitatively assess the risk factors, lifestyles, and contexts facilitating syphilis transmission among heterosexually-identifying individuals in England through the NEXUS study.

**Methods and Findings:** NEXUS was a qualitative study based on semi-structured one-to-one interviews conducted between December 2023 and September 2024. Heterosexually-identifying individuals diagnosed with primary, secondary, or early latent syphilis in the previous year at a sexual health service in one of three regions in England were recruited. Interviews were also conducted with healthcare professionals involved in syphilis management. An analytical framework approach identified salient themes from interview transcripts. The service-user data were triangulated using healthcare professional data.

Interviews were conducted with 19 service-users and seven healthcare professionals. Syphilis acquisition was primarily associated with condomless vaginal or oral sex. Half (10/19) reported multiple partners around the time of diagnosis; other known risk factors, like swinging (multiple concurrent partners) (1/19), sex work (3/19), or transmission among heterosexually-identifying men who have sex with men (1/19), were linked to a minority of infections. Of those reporting a single partner (9/19), four were in a believed exclusive relationship. Many had low perceptions of STI risk but intended to alter sexual behaviours post-diagnosis. Syphilis knowledge was limited among heterosexual individuals and initially seeking medical advice for symptoms from non-sexual health specialists sometimes led to diagnosis delays. Service-users suggested information on syphilis epidemiology, symptoms, and prevention, conveyed through National Health Service sources and mass media, would have been beneficial.

**Conclusions:** We identified a range of contexts associated with syphilis transmission. While multiple partners were common, infections also occurred through infrequent or novel practices or among those with few sexual encounters. Increasing awareness of syphilis among heterosexually-identifying individuals and healthcare professionals working in specialties other than sexual health is important to improve detection, facilitate treatment, and reduce transmission.

## Introduction

Syphilis is a bacterial infection caused by *Treponema pallidum* and is transmitted through sexual contact, as well as vertically (1). Staging based on clinical presentation includes primary, secondary, early latent (when diagnosed within the first two years of infection), late latent, and tertiary (gummatous, cardiovascular, and neurosyphilis); the first three stages are the most infectious and represent early syphilis (2). Syphilis can be effectively treated with antibiotics but can result in chronic and serious complications if left untreated.

The number of infectious syphilis diagnoses in England has increased by 184% over the past decade, from 3,345 in 2013 to 9,513 in 2023 (3). Most diagnoses are among gay, bisexual, and other men who have sex with men (GBMSM) (76% of diagnoses in 2023 (3)), but there have been larger proportional increases among heterosexual men and women in recent years. There were increases of 65% (499 to 825) in women who have sex with men and 33% (851 to 1,133) in men who have sex with women between 2021 and 2023, compared to a 28% increase (5,082 to 6,527) in GBMSM over the same period (4). Combined with evidence of an increasing trend in congenital syphilis diagnoses between 2015 (0.002 cases per 1,000 births) and 2022 (0.014 cases per 1,000 births) (5), this is causing public health concern.

The increase in syphilis diagnoses among heterosexual individuals, also observed across other nations of the UK and Europe (6–9), may be explained by several factors. Previous studies have linked sexual practices such as swinging or group sex to an increased incidence of sexually transmitted infections (STIs) (10, 11), and sex work can also be a risk factor for syphilis (12). A potential key population for syphilis prevention is heterosexually-identifying men who have sex with men (HI-MSM), as they may contribute to STI transmission between MSM and heterosexual networks (13). A greater understanding of the role these factors play in the current syphilis epidemiology in England is needed to contextualise which sexual networks are important for syphilis transmission among heterosexual individuals. It is also important to assess the knowledge and understanding of syphilis in the heterosexual population to recommend appropriate health promotion messages. This information is necessary to inform prevention measures, clinical practice, and public health policy to reduce infection. The NEXUS study aimed to understand the risk factors, lifestyles, and contexts facilitating syphilis transmission among people in England who identify as heterosexual.

## Methods

### Study overview

NEXUS was a qualitative study based on semi-structured one-to-one interviews conducted between December 2023 and September 2024. Participants were service-users attending sexual health services (SHSs) in South Tyneside and Sunderland National Health Service (NHS) Foundation Trust (North East of England), Cambridgeshire Community Services NHS Trust (East of England), and Croydon Health Services NHS Trust (London). Interviews were also conducted with healthcare professionals (HCPs) from participating SHSs and those involved in the writing of national syphilis clinical management guidelines (14). In England, sexual healthcare is provided through an open-access network of SHSs which are free at the point of delivery. These services are staffed by HCPs trained in genitourinary medicine, a medical speciality in the UK which includes the prevention and management of STIs and HIV.

### Participants and recruitment

Service-users were eligible to participate if they identified as heterosexual; were 18 years of age or older; were diagnosed with primary, secondary, or early latent syphilis in the previous 12 months at a participating SHS (the 12 month timeframe was applied from the date of first identifying eligible service-users and did not include the time between identification and interview); understood English sufficiently to arrange an interview (but were offered an interpreter, if needed); and were able to provide informed consent.

Eligible SHS attendees were told about the study by their HCP during a confirmatory, treatment, or follow-up appointment. The depersonalised patient identification codes of eligible service-users in STI surveillance data (15) reported by the study sites were shared with the participating HCPs; additional eligible service-users were identified by clinical staff at the participating SHSs using their electronic patient records management system. HCPs, who could access service-user contact details, explained the study and recorded whether service-users gave verbal permission to be contacted by the researcher in a service log. Service-users who agreed to being contacted were provided with an information leaflet and sheet for review, available in English, Romanian, Lithuanian, Spanish, Portuguese, Bulgarian, and French (these languages were selected based on the distribution of country of birth of service users diagnosed with syphilis in the three study areas). Within a week, contact details (name and phone number) were shared with the researcher, who made up to three attempts to contact each service-user. Recruited service-users were given a unique study ID and invited to an interview, being offered an interpreter if required. A contact log recorded contact details, contact attempts, whether service-users agreed or declined to participate, and where applicable, reasons for declining.

To recruit HCPs, study site principal investigators nominated SHS staff who had experience of working with service-users diagnosed with early syphilis in the last 12 months. A HCP who was also an expert from the British Association for Sexual Health and HIV involved in syphilis clinical management guidelines was selected by the study team. These participants were asked to review a HCP participant information sheet and study summary and invited to an interview. Written or audio recorded verbal consent (as preferred) was provided by all participants (both service-users and HCPs); written consent was required for service-users who requested an interpreter.

## Data collection

Data were collected via audio or video interviews conducted by a trained and experienced interviewer using Microsoft Teams. Participants were offered the choice of audio or video interviews; 16 service-user interviews chose audio and three chose video. All HCP interviews were conducted via video. Service-user interviews lasted 60–90 minutes and HCP interviews lasted 30–40 minutes. One service-user interview required an interpreter. Service-users received a £40 gift card after interview. Interviews followed a semi-structured topic guide; an abridged topic guide was used for the interview involving interpretation. The service-user guide covered relationships, STIs, and sexual behaviour; the experience of syphilis testing and diagnosis; lifestyle and social contexts (in interviews conducted in English only); and communication and information needs related to syphilis. The HCP topic guide focused on testing, diagnosis, treatment, and prevention of syphilis among heterosexual individuals.

## Data analysis

Interviews were audio recorded and transcribed by Microsoft Teams and the interview requiring translation to English was interpreted by a professional agency who signed a confidentiality agreement. All transcripts were anonymised and checked for accuracy using the recordings. An analytical ‘framework’ approach (16, 17) derived initial deductive and inductive codes from the topic guide and three service-user transcripts, chosen to represent different service-user characteristics. The agreed coding index was applied to interview transcripts using NVivo R1 (2020). The data were charted into the framework matrix, using participant terminology where possible, summarised, and interpreted. The service-user data were triangulated using HCP data by identifying comparable and contrasting themes between the two participant groups.

### Ethics

This study was reviewed and approved by the London – Bromley Research Ethics Committee (Reference 23/LO/0699). Participants could withdraw from the study at any time. The study protocol was reviewed by the Health Protection Research Unit in Blood Borne and Sexually Transmitted Infections, sexual health clinicians, and British Association for Sexual Health and HIV members. We sought patient and public involvement and engagement (PPIE) advice from sexual health community stakeholders on the protocol and recruitment materials. Suggestions were incorporated into the study design, including offering interpretation and ensuring the topic guide was culturally sensitive.

## Results

### Characteristics of study participants

Interviews were conducted with 19 service-users and seven HCPs. Amongst service-users who expressed initial interest and agreed to share contact details with study researchers (46), 19 (41%) were interviewed and 27 (59%) were not, a majority 17 (37%) of whom did not respond to contact, five (11%) agreed to interview but did not attend or respond to further follow up, four (9%) could not participate due to other commitments and one (2%) was not interested in participating. Data saturation (where no new themes, insights, or patterns were likely to emerge (16)) was not reached by the end of the study period. Service-users were recruited from South Tyneside and Sunderland (Gateshead and South Tyneside SHSs) (n=9), Cambridgeshire (Bedford and Peterborough SHSs) (n=8), and Croydon (Croydon SHS) (n=2) NHS trusts. All service-users identified as heterosexual (n=9) or straight (n=10), but one male described himself as *‘probably a little bit curious but more straight*’ (P02), referring to some same-sex attraction. Further demographic information is outlined in Table 1. The time between syphilis diagnosis and interview varied between two weeks and 18 months (mean and median time: 7 months), and seven service-users had been diagnosed within the preceding three months. Service-user interviews lasted between 52 and 98 minutes (mean: 74 minutes; median: 72 minutes).

**Table 1:** Service-user participant characteristics.

|  | n (N=19) | % |
| --- | --- | --- |
| **Gender identity** |  |  |
| Men | 10 | 53 |
| Women | 9 | 47 |
| **Age, in years** |  |  |
| 18 to 24 | 5 | 26 |
| 25 to 34 | 4 | 21 |
| 35 to 44 | 5 | 26 |
| 45 and over | 5 | 26 |
| **Ethnic identity<sup>a</sup>** |  |  |
| White British | 14 | 74 |
| White Other | 2 | 11 |
| All other ethnic groups | 3 | 16 |
| **Born in UK/English as first language** |  |  |
| No | 5 | 26 |
| Yes | 14 | 74 |
| **Employment status** |  |  |
| Employed full time | 8 | 42 |
| Employed part time/Student/Retired | 7 | 37 |
| Unemployed | 4 | 21 |
| **Currently struggling financially** |  |  |
| No | 13 | 68 |
| Yes | 5 | 26 |
| Not answered | 1 | 5 |
| **Mental health condition<sup>b</sup>** |  |  |
| No | 11 | 53 |
| Yes | 6 | 37 |
| Not answered | 2 | 11 |
| **Previous STI infection** |  |  |
| No | 13 | 68 |
| Yes | 6 | 32 |
| **Concurrent STI and syphilis diagnosis<sup>c</sup>** |  |  |
| No | 15 | 79 |
| Yes | 4 | 21 |

<sup>a</sup>All other ethnic groups include those of Asian ethnicity, black ethnicity, and mixed ethnicity (England's Census ethnic categories). This data was aggregated into one group to minimise the risk of deductive disclosure due to small numbers. 'White Other' refers to people of White ethnicity that are neither British nor Irish.
<sup>b</sup>Mental health conditions included depression, anxiety, PTSD, and bipolar disorder.
<sup>c</sup>Concurrent STI diagnoses included HIV, herpes, chlamydia, and gonorrhoea.

### Likely route of syphilis acquisition

Service-users were largely confident as to how they acquired syphilis. For most (14/19) respondents this confidence was due to having had few sexual encounters or partners and the timing of partnerships: ‘*I hadn’t had sex like for a year before that*’ (P13). A HCP reflected that some heterosexuals with syphilis likely would never have expected to be diagnosed with an STI.

A quarter (4/19) of all service-users attributed acquiring syphilis to someone they thought was exclusive: ‘*I’ve gone to get a new boyfriend who’s decided to cheat on [me] and just lumber [me] with that now […] because obviously he’s been having sex with other people behind my back and he’s given it to me*’ (P01). Several (5/19) respondents believed they had acquired syphilis from a casual (one-off sexual encounter) partner and 6/19 believed it was acquired from a regular (more than one sexual encounter) partner they were not in an exclusive relationship with; they knew or suspected this partner had had more partners: ‘*in total he’s had like 100 sexual partners*’ (P23) or were ‘higher risk’: ‘*I wouldn’t be surprised if he had sold his services to a woman, not him, but him being paid*’ (P22). Three service-users believed they had acquired syphilis from sex workers (one encounter was outside the UK). One male service-user believed he acquired syphilis from having sex with a male and female couple met through a swingers website.

Around the time of diagnosis half (9/19) of service-users reported one sexual partner, five described two to four partners, and five reported more than four partners. While most (7/9) of those with a single partner attributed their infection to a regular partner, fewer (3/10) with multiple partners did so. Generally, sexual partners were most often met in commercial social venues, such as bars and clubs, and through dating apps, with some being introduced through friends/family.

In thinking about the partner/s from whom they may have acquired syphilis, service-users could reflect on and report their behaviour for a one-off casual partner. However, pinpointing an occasion when syphilis was acquired was more difficult for those with regular partners with whom they had had sex on multiple occasions. The sexual practices they reported with these suspected acquisition partners almost universally included vaginal intercourse (18/19), and commonly oral sex (16/19), but more rarely anal intercourse (4/19); both receptive and active anal intercourse were reported by the male service-user who had sex with a male.

Some HCPs suggested that although sometimes they diagnosed syphilis in heterosexually-identifying men who had sex with men (HI-MSM), they had seen a shift over the last decade towards increased diagnoses in men with exclusively female partners. Only two service-users reported condom use for vaginal or anal sex (both one-off casual partners), although none reported their use for oral sex: *‘I think it was only when it was penetration’* (P02). A few service-users engaged in sexual practices with the partner they believed they acquired syphilis from that were irregular for them, such as their first experience of oral sex without a condom, or anal intercourse: *‘it was the only […] sexual partner who was performing oral sex without protection, without a condom’* (P15).

### Perceptions of sexual and reproductive health risk

Service-users’ perceptions of sexual risk, personally to them and generally, varied. A few (4/19) did not actively consider STI risk because they did not see or were not interested in information about it: ‘*I never really hear about them. So I just kind of like, forget they’re there*’ (P13). Some (5/19) trusted their partners to keep relationships exclusive or to inform them of infection, or they assumed that having few sexual partners minimised their risk. While a similar proportion had previously considered risks related to pregnancy (13/19) and STIs (12/19), reporting a sole or greater consideration for pregnancy risk (8/19) was more common than STI risk (2/19). Four service-users reported that they did not have much consideration for either, and a few female service-users using oral contraception relied on their partners to manage condom use: ‘*Yeah it was more contraception. […] It didn’t cross my mind, STIs. I thought everyone was clean. I thought everyone would tell the truth*’ (P04). Some (8/19) occasionally practiced condom use in situations where STI transmission risk was perceived as greater, so with new, single encounter, or paid for sex partners. Half (10/19) discussed the possibility of syphilis transmission either through contact with genital ulcers, though condomless oral sex, or condom failure: ‘*Well, condoms can’t protect you from anything, and they can break*’ (P26), *‘I thought you got less chance of catching it off oral but obviously I did’* (P02).

Substance use, predominantly alcohol, was reported to play a significant role in half (10/19) of the service-users’ sexual decision-making. A few individuals acknowledged making riskier choices when under the influence, including having condomless sex and partners they would not otherwise choose: ‘*I think if I was sober I would never do that’* (P13).

Following their syphilis diagnosis, which was the first ever STI diagnosis for most (13/19) service-users, all expressed consideration for STIs: *‘I think I’m obviously just more aware now, I think I was very unaware of things that you could get through sex’* (P12), and many (14/19) expressed a desire to alter their sexual behaviours (the 14 individuals comprised 7 women and 7 men). The majority (15/19) stated they would be more cautious in selecting partners, either by choosing different types of partners (e.g., not one-off, swinger, or sex worker); abstaining (2/19) or reducing partner numbers (11/19); and introducing, or being more consistent in, using condoms, particularly with new partners (12/19): ‘*Before I just, I didn’t really care. […] Like I never really used a condom. But now I think I’ve learned my lesson*’ (P23). While many had attributed riskier behaviours to alcohol use, two service-users expressed an intention to avoid similar situations in the future: ‘*I’m going to be so much more careful when I’m drunk. Like I don’t want to get in that situation again*’ (P13). A few (4/19) service-users planned to engage in more regular STI testing between partners.

### Patient journey

The most common reason given for testing for syphilis was having, and wanting relief from, symptoms (14/19 service-users, Table 2). Many (12/19) service-users suspected they had a STI before they had tested, but only five suspected they had syphilis; suspicions of syphilis mainly came from researching symptoms: ‘*the sore it wasn’t going away, it was getting bigger and so I thought right, I’ll Google it, and syphilis came up straight away’* (P04). Genital symptoms were described as uncomfortable and somewhat painful by men, but much more painful by women: ‘*it burns and stings […] like the way I describe it is if you’ve got children or if you’re in labour and you get that ring of fire when you’re trying to push his head out*’ (P01). It should be noted that symptoms may also have been a result of concurrent STIs in the case of four service-users. Most service-users sought help between two days and three weeks after noticing symptoms.

**Table 2.** Reasons for testing for syphilis by the initial service used and whether service-users suspected they had a STI.

| n (N=19) | Reason for testing |  |  |  |  |  |  |
| --- | --- | --- | --- | --- | --- | --- | --- |
|  | Symptoms |  | General check-up |  | Partner notification |  |  |
| Initial service | Suspected STI | Did not suspect STI | Suspected STI | Did not suspect STI | Suspected STI | Did not suspect STI | Total |
| Sexual health service | 7 | 0 | 0 | 1 | 2 | 1 | 11 |
| Online postal self-sampling kit | 1 | 0 | 0 | 1 | 0 | 0 | 2 |
| General practitioner (GP) | 2 | 3 | 0 | 0 | 0 | 0 | 5 |
| Dental hospital | 0 | 1 | 0 | 0 | 0 | 0 | 1 |
| **Total** | 10 | 4 | 0 | 2 | 2 | 1 | 19 |

The minority who did not suspect an STI said that they had sought help from non-STI clinicians (including general practitioners (GPs), a dentist, an ophthalmologist, and an ear, nose, and throat (ENT) consultant) for non-genital symptoms, such as diffuse rash, mouth ulcers, eyesight problems, swollen lymph nodes, or neurological issues. This sometimes led to a delayed syphilis diagnosis: ‘‘*don’t worry, we will get to the bottom of it’, he said. ‘I’m ruling everything out that I can think of’ and eventually they came up with syphilis*’ (P22). HCPs noted gaps in the ability of some non-STI specialists to identify syphilis and described the importance of increasing awareness among HCPs to test people with different presentations due to syphilis ‘*being the great mimicker*’ (HCP06). Universal testing for syphilis was suggested to identify infections that would not otherwise be recognised: ‘W*e’d get a steady stream of diagnoses in those groups of people who have had unexplained presentations, sometimes for over a year and they’ve seen multiple, you know, specialists who’ve been unable to make a diagnosis. And eventually someone does a syphilis blood test’* (HCP01).

Two interviewees had a general check-up; one was asymptomatic and tested via online postal self-sampling because ‘*I find the clinic just kind of intimidates me*’ (P12). The other sought help for bleeding and discomfort during sex. Three service-users tested due to partner notification. One was asymptomatic, but the others attributed symptoms to allergies (rash), having dry sex (lacerations on penis), and a broken tooth (mouth sores): ‘*I thought there is no point getting tested like I’m alright, I haven’t got anything*’ (P39).

Several (12/19) service-users described their syphilis diagnosis as being shocking and embarrassing: ‘*I think there’s such a stigma around like STIs […] like people think ‘oh because you’ve got one you must be like sleeping around with loads of boys’*’ (P13). Most people did not share their diagnosis with anyone, as there was fear of judgement: ‘*it’s not something you want to go shouting from the rooftops is it?*’ (P22). Those who did share their diagnosis suggested people they told were also surprised but supportive.

Many service-users were focused on treatment, which meant some struggled to attend to or recall information provided: ‘*I was like ‘I don’t really care what it is, I just need the treatment and I want it to be done as soon as possible’*’ (P23). The most common treatment was penicillin injections (14/19 service-users), then oral (4/19) or intravenous (1/19) antibiotics. Three service-users were given treatment before syphilis confirmation, one of whom remained unaware of their diagnosis until they returned for testing a year later: ‘*I wasn’t aware for the whole year that that’s actually what I’d contracted. I don’t know if I did know and I’d forgotten, but I felt like that was the first time I found out*’ (P28). One HCP related other examples of this: *‘It might be that they had that explained to them a long time ago, and they’ve forgotten, […] they didn’t quite grasp it at the time, or it wasn’t explained properly at the start.’* (HCP06).

Support with partner notification was offered to all service-users, often simultaneously with treatment ‘*so that nobody’s missed, because once you let them out of the clinic, we don’t know whether we will be able to contact them or not*’ (HCP03). Some service-users only contacted their most recent partners or those they suspected they acquired syphilis from. All service-users were amenable to follow-up syphilis testing, but two service-users had not yet been contacted by the SHS to make an appointment. HCPs reported varying success with follow-up: ‘*once they know they’re not infected then they’ll stop coming, but initially they do come back*’ (HCP03), ‘*I had many patients who did not come back for follow up. And this is a big problem.*’ (HCP05).

### Knowledge and understanding of syphilis testing, treatment, and prevention

Prior to their diagnosis, most (17/19) service-users had little to no knowledge of syphilis, with 4/19 having never heard of it: ‘*I was upset, but […] I didn’t really know what syphilis was’* (P12). HCPs observed this limited awareness, suggesting a requirement for greater explanation: ‘*You’re using this word and term that no one’s ever heard of. It’s not really in the public, certainly not in the heterosexual public eye’* (HCP04). Limited experience and understanding of STIs influenced first perceptions that syphilis was akin to HIV, with service-users expressing primary concerns about treatment and severity. Some contrasted syphilis with more familiar STIs like chlamydia or gonorrhoea, believed to be less severe and more easily treatable: *’I just knew it was a serious STI really […] I never really paid much thought to it. Because you know, everyone talks about like, chlamydia, gonorrhoea, they’re all very treatable, very common, but like no one ever really speaks about syphilis or HIV*’ (P23).

While 15 service-users had heard of syphilis, their understanding was vague, with many viewing it as an archaic or rare disease. Most (10/19) service-users recognised syphilis as potentially severe, though their understanding was often imprecise, associating it with historical deaths rather than specific symptoms: ‘*I’d heard of it, but I didn’t really know anything about it […] I didn’t know any of the symptoms or anything’* (P03). Only three service-users knew syphilis was a bacterium or were aware of its treatment options.

Information sources varied, with many service-users having sought information after developing symptoms or between testing or diagnosis and treatment, turning to online information such as Google searches or highly trusted NHS information. Some service-users discussed the provision of verbal and/or written information by HCPs, and this was reported to be trusted and valued, particularly regarding treatment and reassurance: ‘*He reassured me that the medication would take it away and I trusted him, and it all worked out’ (P04).* Social media online chat groups and informal networks, such as family or friends, also provided information for a few service-users: *‘I was like on every website, on Reddit, I was just like scrolling for hours, researching about it’* (P23). A few service-users gained limited knowledge of syphilis through film, television, or previous sex education. Most service-users expressed a post-hoc desire for information on syphilis before diagnosis, through sources such as mass media, community forums, and online symptom searchers. Potentially useful information would have regarded the existence or prevalence of syphilis, signposting the potential for infection*: ‘to know that it can happen to anybody. Don’t just think ‘Oh, it’ll never be me’ because it could be, you know, and it was’* (P02). However, some suggested they would not have attended to any information: ‘*I don’t think there’s any information that […] could be given before that would, you know, change the way I acted […] So it doesn’t seem particularly helpful to try’* (P20), hence HCPs emphasised the importance of regular testing. Some service-users (4/19) noted that greater awareness of syphilis might have led to earlier diagnosis through symptom recognition but would not necessarily have changed their sexual behaviours: *‘well it wouldn’t have probably stopped us getting it, but it would have made us get a check quicker’* (P18).

Despite treatment, a minority (5/19) had some uncertainty about subsequent testing in terms of whether future test results indicated active infection or past exposure: *‘Now if he says to me, activity level, it means there’s still something there’* (P22). Almost all (18/19) understood that a complete cure was possible, and half did not realise that it can take 12 weeks for first symptoms to show. The understanding of infectiousness varied, with some believing a two-, four-, or six-week period post-treatment was sufficient before resuming sexual activity ‘*So they said I can’t have sex for two weeks and that finishes today’* (P23), whereas others suggested they should wait until further testing or were uncertain of the waiting period: ‘*That’s what I’m a little bit confused [about], even if I’m negative then […] I have to use the condoms’* (P30). HCPs shared good practices for ensuring understanding of the syphilis diagnosis, infectiousness, and future testing such as using analogous explanations and drawing comparisons to diseases like chickenpox.

## Discussion

We conducted a qualitative study of service-users and HCPs and found that syphilis transmission among people identifying as heterosexual in England was linked to a combination of factors and contexts. Syphilis acquisition was primarily associated with condomless vaginal or oral sex. Half of the interviewed service-users reported multiple partners around the time of diagnosis, but engagement with other known syphilis risk factors, such as swinging (multiple concurrent partners), sex work, or sex between men, was linked to a minority of infections. We found that syphilis acquisition was often unexpected due to having few sexual encounters, perceived exclusivity, or engaging in their first occasion of an irregular sexual practice. Syphilis knowledge was very limited, and syphilis was often perceived as rare, leading to feelings of shock when diagnosed, particularly when service-users presented with non-genital symptoms and diagnoses were delayed. Compared to other bacterial STIs, there was less knowledge and a greater degree of stigma associated with syphilis. Few service-users had prioritised STI prevention, but most intended to change this post-diagnosis, and suggested they would have benefitted from sexual health promotion messages on syphilis prevalence, symptoms, treatment, and prevention.

This study was designed and conducted by a multidisciplinary team, with input from researchers, clinicians, and community-based organisations, and investigated the experiences and perceptions of both service-users and expert HCPs. Service-users were identified and recruited from multiple SHSs in three study areas, ensuring a geographic spread and breadth of experience. The experiences of those tested and treated through SHSs in other areas or non-SHSs only, and those who did not have syphilis infection identified were not included. Those service-users who agreed to take part in the study may be more engaged with their syphilis diagnosis, may perceive it as less stigmatising, may be more willing to discuss and seek to understand their infection, and may be more likely to have been approached soon after diagnosis and initial treatment. They may not necessarily represent the heterogeneous characteristics and experiences of heterosexually-identifying people with syphilis in England. A larger study recruiting more service-users may provide further insights into the range of factors associated with syphilis transmission in heterosexual individuals, given the heterogeneity of experience found in the NEXUS study, which likely did not enable data saturation to be reached. It could also be recommended for a larger study to use purposive sampling to ensure representation of minoritised ethnic groups and non-UK born service-users.

While the experience of those diagnosed more than a year previously was not sought, the seven-month average time since diagnosis may result in some recall bias arising from forgetting or changes to thinking as a response to greater reflection. However, this time lag reflects the challenges faced in recruiting service-users diagnosed with an acute infection, many of whom may wish to put the episode behind them, in contrast to recruiting people diagnosed with a chronic condition. These challenges were reported by recruiting SHSs and have also been recognised in PPIE in sexual health research (18).

Following PPIE input to enhance inclusion, study materials were provided in six additional non-English languages reflecting common countries of origin among potential service-users, and interpretation services were also offered but only used in one interview. We used semi-structured interviews, which enable in-depth investigation of previously identified salient questions and options for participants to discuss aspects not initially considered. Analysis followed the ‘framework’ method, allowing a systematic and flexible analysis approach (17), which was helpful in data organisation and in allowing comparisons to be drawn between participant data.

We found that the behaviour most commonly associated with syphilis acquisition was having multiple sexual partners, which was the case for half of our service-user sample. A previous heterosexual syphilis outbreak in the US was attributed to having multiple sexual partners coinciding with drug use (19). Another emergent theme from our analysis was unexpected infection acquisition given partnership circumstances. Specifically, many service-users had few sexual encounters or partners; presumed exclusivity; engaged in their first occasion of an irregular sexual behaviour for them, such as condomless sex; or chose a partner who may have been perceived as being at greater risk of STIs in contrast to the individual’s previous partners, such as casual one-off partners, swingers, or sex workers. Individuals who were in a believed monogamous relationship did not consider themselves to be at risk, and in other cases, having a low sexual risk perception may have facilitated syphilis transmission. For example, many interviewees reported little prioritisation or consideration of STI risk and greater consideration for pregnancy risk, which are common findings among heterosexual individuals (20, 21). A nationally representative British survey revealed low levels of perceived STI risk among mostly heterosexual men and women, even among those engaging in condomless sex with new/multiple partners in the previous year (22). Condomless sex was universally linked to the likely route of syphilis acquisition, even though condom use was sometimes reported in previous encounters with perceived higher risk partners. Although some saw condoms as protective, half of service-users interviewed questioned their effectiveness generally and in the context of syphilis transmission. Health promotion messaging should emphasise condom use (including use for oral sex and clarifying how to use effectively) to prevent syphilis transmission (23).

There were few examples of service-users or their partners having same-sex encounters and HCPs reported diagnosing more syphilis in behaviourally heterosexual men than in HI-MSM, suggesting sex between men was not a key driver of heterosexual syphilis in our study population. We only gained insight into individual circumstances and views of HCPs in limited geographical areas, so HI-MSM may play a role in heterosexual syphilis transmission, given evidence of some mixing between GBMSM and heterosexual networks (24) in other areas of England.

It has also been proposed that substance use may contribute to increasing heterosexual syphilis cases (24); a previous study examining sexualised drug use among heterosexual individuals found associations between substance use, including alcohol, and having more sexual partners, engaging in condomless sex, and being diagnosed with syphilis (25). These findings were based on a large cross-sectional survey and correspond to our findings on alcohol use-driven riskier sexual decision-making reported by some of the service-users interviewed. However, only a minority attributed acquiring syphilis to substance use and more considered its impact on their sexual behaviour generally.

Our findings showed that a syphilis diagnosis was often unexpected by service-users and non-sexual health HCPs alike. There was a lack of awareness of the potential for syphilis acquisition and symptoms, with the presentation of genital symptoms often suspected to be other STIs such as herpes (although only a minority of service-users were diagnosed with a concurrent STI), and non-genital symptoms attributed to a range of other health issues including allergies and oral health conditions. Increased symptom recognition may have led to earlier diagnosis as some service-users dismissed symptoms and did not seek healthcare until notified by partners who had been diagnosed with syphilis. Delaying health service seeking despite STI symptom presentation has been previously observed among men (26), hence interventions aimed at raising awareness of the importance of symptom recognition among heterosexual individuals could encourage prompter testing and contribute to syphilis control efforts.

Syphilis testing was often not directly sought, with many service-users initially seeking information after developing symptoms and some attending non-sexual health services. A previous mixed-methods study found that men and women in England prioritised emotional reassurance from others’ lived experience and presenting symptoms to a GP over accessing medical expertise at SHSs (27). When non-sexual health clinicians were consulted, other investigations were sometimes conducted before a syphilis test was suggested, leading to diagnosis delays. As suggested by HCP interviewees and acknowledged in England’s national Syphilis Action Plan (13), it is therefore important to recognise and improve the awareness of syphilis epidemiology and presentation among non-STI specialist HCPs. The provision of accurate and accessible information on syphilis to both non-sexual health HCPs and the heterosexual population may help expedite attendance at SHSs (27) and ensure that people access appropriate and timely testing and healthcare. Social media and mass media were suggested as potentially useful sources for syphilis health promotion among the heterosexual population (28). The use of a swingers website to meet partners reported by one service-user demonstrates a potential opportunity for syphilis health promotion online, targeting users of such websites. Education which highlights aspects such as syphilis epidemiology and rising infection rates, ease of testing, and highly effective treatment to avoid severe health consequences may minimise stigma (29). However, consideration needs to be given to those who said they would not have attended to STI messaging or changed sexual behaviour, as risk perception may not be necessary or sufficient for seeking STI care (22). Expanding syphilis testing to universal opt-out screening in emergency departments may help to detect infections that would not otherwise be identified (30–32).

There were some outstanding knowledge needs post-diagnosis, including an example of a service-user being unaware of their syphilis diagnosis and treatment until they returned to a SHS a year later. There was some uncertainty regarding infectiousness and prevention post-treatment. Improving understanding of the syphilis diagnosis, subsequent testing, treatment efficacy, and future prevention should therefore be considered; good practice examples, such as using comparisons between syphilis and chickenpox as noted by a HCP, could be shared. It may be beneficial for HCPs to ensure information on these aspects are conveyed during face-to-face appointments or in the provision of written information (with population health literacy levels considered (33)), as service-users reported these to be trusted sources.

We found that a syphilis diagnosis often prompted intentions to implement behaviour change to prevent future STIs, including consistent condom use and choosing partners with a perceived lower risk of STIs. These findings align with other research where sexual-risk-reduction behaviours were considered following a new STI diagnosis, and a 3-month follow up of this cohort suggested these intentions were successfully acted upon (34). Focusing on risk-reduction counselling for individuals who do not intend to change behaviour could be a priority to prevent reinfection, which is an important element of syphilis control (13, 34). To enable this, it is important to consider the capacity and demand of SHSs and the value of maintaining skilled sexual health advisors (HCPs with expertise in partner notification for STIs) to deliver such services.

In conclusion, the NEXUS study identified a range of contexts surrounding syphilis transmission among those who identify as heterosexual, with having multiple partners being a common behaviour associated with acquisition. Many individuals had low STI risk perceptions and did not prioritise prevention, which may have facilitated transmission. We found a knowledge gap in the heterosexual population and considered the importance of increasing syphilis awareness for potential impacts on stigma reduction and identifying diagnoses that may be missed. Syphilis health promotion could utilise materials provided in health services, as these were considered a trusted information source, and mass media to reach a wider audience. It is also important to promote syphilis education among non-sexual health HCPs, as we found that help-seeking outside of SHSs could lead to diagnosis delays. This highlights the need for multi-agency working across the health system and could be explored further in future studies focusing on the syphilis education needs of non-sexual health HCPs. Overall, these findings emphasise that health promotion efforts among heterosexual populations and increasing knowledge of syphilis among non-sexual health HCPs are necessary to raise awareness of syphilis with the aim of improving detection, facilitating treatment, and reducing transmission.

## Data Availability

The data that support the findings of this study have been assessed by the UK Health Security Agency’s Office for Data Acquisition and Release as having sensitive personal information and are therefore not publicly available to protect participant privacy.

## Abbreviations

GBMSM: gay, bisexual, and other men who have sex with men
HCP: healthcare professional
HI-MSM: heterosexually identifying men who have sex with men
HPRU: Health Protection Research Unit
NHS: National Health Service
NIHR: National Institute for Health and Care Research
PPIE: patient and public involvement and engagement
SHS: sexual health service
STI: sexually transmitted infection

## Acknowledgements

We would like to thank all staff at the participating sexual health services that enabled recruitment, and all participants of the NEXUS study. Thank you to the Terrence Higgins Trust who gave PPIE feedback. We additionally thank the reviewers of the study protocol: Margaret Kingston, Raquel Bosó Pérez, Jo Kesten, Shema Tariq, and Lorraine McDonough.

We acknowledge members of the National Institute for Health and Care Research Health Protection Research Unit (NIHR HPRU) in Blood Borne and Sexually Transmitted Infections (BBSTI) Steering Committee: Professor Caroline Sabin (HPRU Director), Dr John Saunders (UKHSA Lead), Professor Catherine Mercer, Professor Gwenda Hughes, Dr Hamish Mohammed, Professor Greta Rait, Dr Ruth Simmons, Professor William Rosenberg, Dr Tamyo Mbisa, Professor Rosalind Raine, Dr Sema Mandal, Dr Rosamund Yu, Dr Samreen Ijaz, Dr Fabiana Lorencatto, Dr Rachel Hunter, Dr Kirsty Foster and Dr Mamooma Tahir.

